# Plasma ACE2 levels predict outcome of COVID-19 in hospitalized patients

**DOI:** 10.1101/2021.03.08.21252819

**Authors:** Tue W. Kragstrup, Helene Søgaard Singh, Ida Grundberg, Ane Langkilde-Lauesen Nielsen, Felice Rivellese, Arnav Mehta, Marcia B. Goldberg, Michael Filbin, Per Qvist, Bo Martin Bibby

## Abstract

**Background:** Severe acute respiratory syndrome coronavirus 2 (SARS-CoV-2) binds to angiotensin converting enzyme 2 (ACE2) enabling entrance of the virus into cells and causing the infection termed coronavirus disease of 2019 (COVID-19). COVID-19 is a disease with a very broad spectrum of clinical manifestations, ranging from asymptomatic and subclinical infection to severe hyperinflammatory syndrome and death.

**Methods:** This study used data from a large longitudinal study of 306 COVID-19 positive patients and 78 COVID-19 negative patients (MGH Emergency Department COVID-19 Cohort with Olink Proteomics). Comprehensive clinical data were collected on this cohort, including 28-day outcomes classified according to the World Health Organization (WHO) COVID-19 outcomes scale. The samples were run on the Olink® Explore 1536 platform which includes measurement of the ACE2 protein.

**Findings:** High baseline levels of ACE2 in plasma from COVID-19 patients were associated with worse WHOmax category at 28 days with OR=0.56, 95%-CI: 0.44-0.71 (*P* < 0.0001). This association was significant in regression models with correction for baseline characteristics, pre-existing medical conditions, and laboratory test results. High levels of ACE2 in plasma from COVID-19 patients were also significantly associated with worse WHO category at the time of blood sampling at both day 0, day 3, and day 7 (*P* = 0.0004, *P* < 0.0001, and *P* < 0.0001, respectively). The levels of ACE2 in plasma from COVID-19 patients with hypertension were significantly higher compared to patients without hypertension (*P* = 0.0045). The plasma ACE2 levels were also significantly higher in COVID-19 patients with pre-existing heart conditions and kidney disease compared with patients without these pre-existing conditions (*P* = 0.0363 and *P* = 0.0303, respectively). There was no difference in plasma ACE2 levels comparing patients with or without pre-existing lung disease, diabetes, or immunosuppressive conditions (*P* = 0.953, *P* = 0.291, and *P* = 0.237, respectively). The associations between high plasma levels of ACE2 and worse WHOmax category during 28 days were more pronounced in COVID-19 positive patients compared with COVID-19 negative patients but the difference was not significant in the two-way ANOVA analysis.

**Interpretation:** This study suggests that measuring ACE2 is potentially valuable in predicting COVID-19 outcomes. Further, ACE2 levels could be a link between severe COVID-19 disease and its risk factors, namely hypertension, pre-existing heart disease and pre-existing kidney disease. The design of the data analysis using the Olink platform does not allow assessment of quantitative differences. However, previous studies have described a positive correlation between plasma ACE2 and ACE1 activity. This is interesting because ACE1 (serum ACE) analysis is a standardized test in most hospital laboratories. Therefore, our study encourages quantitative investigations of both plasma ACE 1 and 2 in COVID-19.

**Key Points:** *Question:* Can plasma levels of the receptor for severe acute respiratory syndrome coronavirus 2 (SARS-CoV-2), angiotensin converting enzyme 2 (ACE2), predict outcome of coronavirus disease of 2019 (COVID-19).

**Findings:** In this study of 306 COVID-19 positive patients, high baseline levels of ACE2 in plasma from COVID-19 patients were associated with worse outcome measured by the World Health Organization (WHO) COVID-19 outcomes scale.

**Meaning:** Measuring ACE2 is potentially valuable in predicting COVID-19 outcomes and link COVID-19 disease and the risk factors hypertension, pre-existing heart disease and pre-existing kidney disease.

## Introduction

Since December 2019, a previously undiscovered virus, severe acute respiratory syndrome coronavirus 2 (SARS-CoV-2), has caused a devastating global pandemic. The disease caused by SARS-CoV-2 infection has been termed coronavirus disease of 2019 (COVID-19) with clinical manifestations ranging from asymptomatic and subclinical infection to severe hyperinflammatory syndrome and death.^1^ Risk factors for fatal infection are male gender, increased age and comorbidities such as pre-existing hypertension, heart disease, lung disease, diabetes, and kidney disease.^2,3^

SARS-CoV-2 binds to the angiotensin converting enzyme 2 (ACE2) receptor enabling entrance into cells through membrane fusion and endocytosis.^4-7^ The ACE2-receptor is distributed in different tissues such as vascular endothelial cells, smooth muscle cells, nasal and oral mucosa, enterocytes within the intestines, and is especially abundant in type II alveolar pneumocytes in the lungs.^8,9^ This distribution explains possible entry routes for the virus, and why target cells such as the pneumocytes are highly vulnerable to viral infection.^9^

ACE2 is part of the renin-angiotensin-aldosterone-system (RAAS). Renin cleaves angiotensinogen leading to formation of angiotensin I (Ang I). Ang I is then converted to angiotensin II (Ang II) through cleavage by angiotensin-converting-enzyme (ACE), which is found in the vascular endothelium and more plentifully in the pulmonary endothelium.^10,11^ ACE2 opposes the effects of the RAAS by cleavage of Ang I and Ang II, thereby attenuating increases in blood pressure.^12^ Dysregulation of RAAS is therefore implicated in many diseases such as hypertension and kidney disease.^11,13^

Plasma ACE2 activity is elevated in patients after COVID-19 infection compared to healthy controls.^14^ Further, ACE2 plasma concentrations have been measured in patients with risk factors for severe COVID-19 disease. In patients with heart failure, plasma levels of ACE2 were higher in men than in women which could explain the higher incidence and fatality rate of COVID-19 in men.^15^ Increased plasma ACE2 concentration have also been associated with increased risk of major cardiovascular events.^16^ Finally, ACE2 levels were recently shown to be significantly elevated in serum from smokers, obese and diabetic individuals.^17^ Therefore, strategies to use soluble recombinant ACE2 as a treatment in COVID-19 are being investigated.^18^

This study is the first description of ACE2 levels in association with disease outcomes in patients with COVID-19 disease. Data were retrieved from a publicly available dataset from a large longitudinal COVID-19 study provided by Olink Proteomics.^19^

## Methods

### Study populations

Detailed description is available online (https://www.olink.com/mgh-covid-study/) (Fig. 1).^19^ The samples were extracted from a study that included patients 18 years or older, who were in acute respiratory distress with a clinical concern for COVID-19 upon arrival to the Emergency Department. Of the 384 patients enrolled, 306 (80%) tested positive and 78 tested negative for SARS-CoV-2. COVID-19-positive patients had blood samples drawn on days 0, 3, and 7, while virus-negative patients only had samples drawn on day 0. Clinical data collected included 28-day outcomes classified according to the World Health Organization (WHO) COVID-19 outcomes scale. 1 = Death. 2 = Intubated, ventilated, survived. 3 = Non-invasive ventilation or high-flow nasal cannula (this treatment is not used in the management of patients with COVID-19 or with suspected COVID-19 and was therefore not part of this study). 4 = Hospitalized, supplementary O_2_ required. 5 = Hospitalized, no supplementary O_2_ required. 6 = Not hospitalized.^20^ Patient age and body mass index (BMI) and pre-existing medical conditions were registered, including heart conditions (coronary artery disease, congestive heart failure, valvular disease), kidney disease (chronic kidney disease, baseline creatinine >1.5, end stage renal disease), lung disease (asthma, chronic obstructive lung disease, requiring home O_2_, any chronic lung condition), diabetes (pre-diabetes, insulin and non-insulin dependent diabetes), and immunosuppressive conditions (active cancer, chemotherapy, transplant, immunosuppressive agents, asplenia). Laboratory tests used in the analyses of this study were C-reactive protein (CRP), absolute neutrophil count, and D-dimer. Continuous variables were categorized to make the dataset anonymized. Age (years) categories were 1=20-34, 2=36-49, 3=50-64, 4 =65-79, 5=80+. BMI (kg/m^2^) categories were 0=<18.5 (underweight), 1=18.5-24.9 (normal), 2=25.0-29.9 (overweight), 3=30.0-39.9 (obese), 4=40+ (severely obese), 5=Unknown (these were excluded from analysis on BMI). CRP (mg/L) categories were 1=0-19.9, 2=20-59.0, 3=60-99.9, 4=100-179, 5=180+. Absolute neutrophile count (10^9^/L) categories were 1=0-0.99, 2=1.0-3.99, 3=4.0-7.99, 4=8.0-11.99, 5=12+. Ddimer (fibrinogen-equivalent units) categories were 1=0-499, 2=500-999, 3=1000-1999, 4=2000-3999, 5=4000+.

**Fig. 1.**
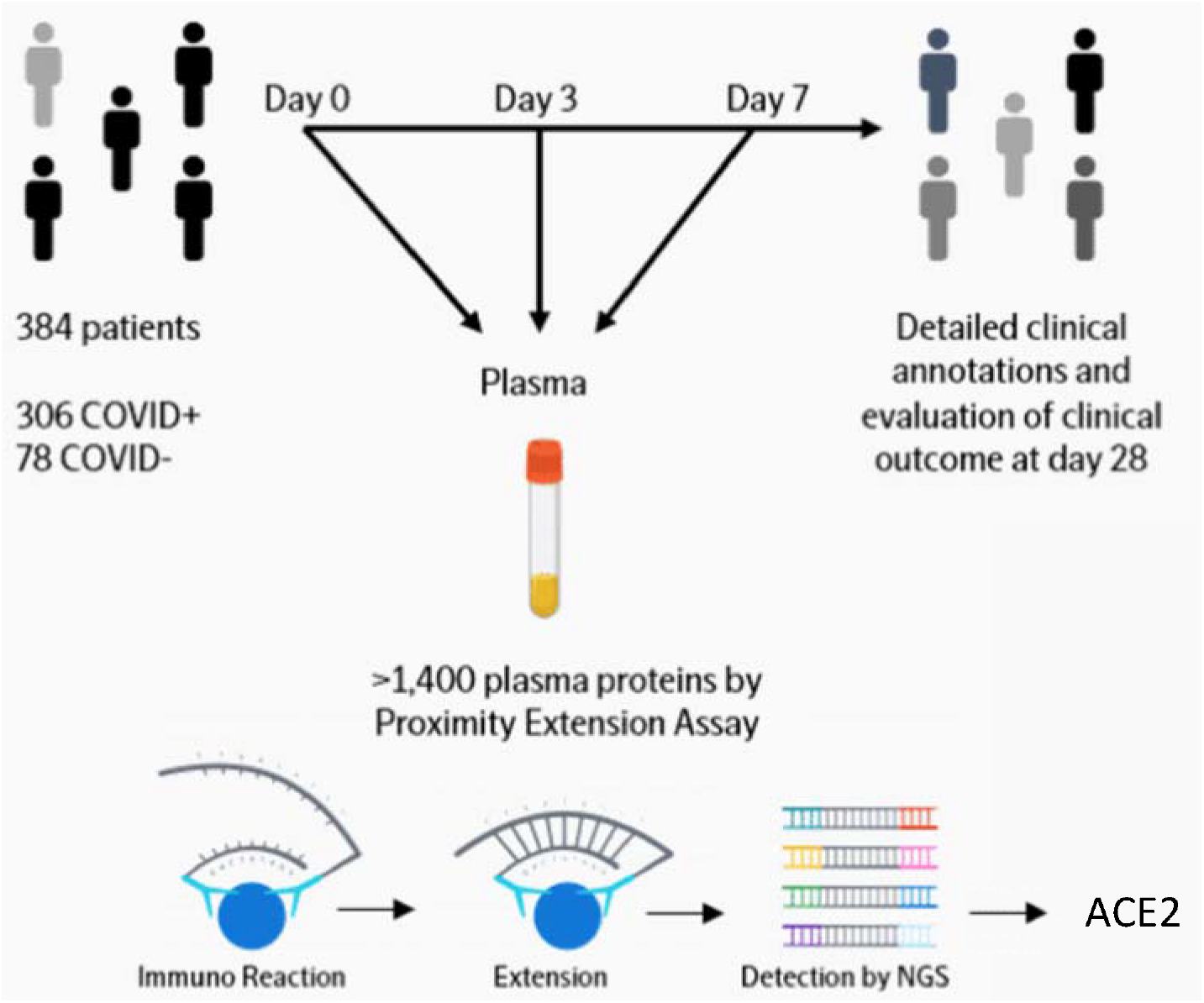
Diagram of study work flow. Link https://www.olink.com/mgh-covid-study/. Modified and used with permission.

### Quantification of ACE2 and Olink data analysis

Detailed description is available online (https://www.olink.com/mgh-covid-study/) (Fig. 1).^19^ Briefly, the samples were analyzed by the Olink® Explore 1536 platform which includes measurement of the ACE2 protein. The Olink platform is based on the Proximity Extension Assay (PEA) technology developed for high-multiplex analysis of proteins in just 1 µL of sample.^21^ In PEA, a matched pair of antibodies, each carrying a unique DNA-tag, bind to the target protein in a sample. Upon binding, the DNA will come in close proximity and hybridize generating a double-stranded barcode used for digital identification, amplification and detection using next-generation sequencing. Olink® Explore 1536 consists of 1472 validated proteins and 48 controls and covers low-abundant inflammation proteins, actively secreted proteins, organ-specific proteins, drug targets, etc. The PEA technology has a built-in quality control system consisting of technical and sample controls to monitor the performance of the assay and the individual samples. Data generation consists of three main steps: normalization to known standard (extension control), log2-transformation, and level adjustment using the plate control. The generated data represent relative protein values, Normalized Protein eXpression (NPX), on a log2 scale where a larger number represents a higher protein level in the sample. For more information about Olink® Explore 1536, PEA and NPX, please visit www.olink.com.

### Statistical analyses

Data were downloaded from https://www.olink.com/mgh-covid-study/. Group comparisons were made using non-parametric tests. The diagnostic value of ACE2 levels at baseline was further tested with a receiver operating characteristic (ROC) curve for WHOmax groups 1-2 (died or intubated during the 28-day period) vs WHOmax groups 4-6 (hospitalized without intubation or not hospitalized). The comparison of WHOmax group differences between COVID-19 positive and negative patients which was done using a two-way ANOVA. The association between WHOmax (response variable) and ACE2 (explanatory variable) was also investigated using ordered logistic regression and results are presented as odds ratios for lower WHOmax scores (worse disease) per unit increase in ACE2. The specific statistical method tests used for each analysis is described in the table and figure legends. In all tests, the level of significance was a two-sided *P* value of less than 0.05. Figures were made using GraphPad Prism 8 for Mac (GraphPad Software).

## Results

### Association between day 0 plasma ACE2 and WHOmax groups in COVID-19 patients

First, we investigated the association between ACE2 levels at day 0 and WHOmax. WHOmax category at 28 days is the maximum WHO score within first 28 days with death being the maximum possible. High baseline levels of ACE2 in plasma from COVID-19 patients were significantly associated with worse WHOmax category at 28 days with OR=0.56, 95%-CI: 0.44-0.71 (P < 0.0001) (Fig. 2 and Table 1). In the receiver operating characteristic (ROC) curve for WHOmax groups 1-2 (died or intubated during the 28-day period) vs WHOmax groups 4-6 (hospitalized without intubation or not hospitalized) the area under the curve (AUC) was 0.67, 95%-CI: 0.60-0.73 (Fig. 2). The association between plasma ACE2 levels at day 0 and WHOmax was also tested in regression models with correction for baseline characteristics, pre-existing medical conditions, and laboratory test results. In models correcting for age, body mass index, hypertension, and pre-existing heart conditions, kidney disease, lung disease, diabetes, and immunosuppressive conditions significant associations were still observed between the plasma ACE2 levels at day 0 and WHOmax (Table 1). Furthermore, we analyzed the association between the plasma ACE2 levels at day 0 and WHOmax after correcting for CRP, absolute neutrophile count, and D-dimer to evaluate whether plasma ACE2 levels adds to the information already achieved by these laboratory test results. The association between high ACE2 and worse WHOmax category remained significant (Table 1).

**Table 1.** Associations between day 0 plasma concentrations of ACE2 and WHOmax group in COVID-19 positive patients inordered logistic regression models with WHOmax as outcome.

| Model | n | OR | 95%-CI | p |
| --- | --- | --- | --- | --- |
| 1 | 285 | 0.56 | 0.44-0.71 | <0.0001 |
| 2 | 285 | 0.54 | 0.42-0.70 | <0.0001 |
| 3 | 285 | 0.54 | 0.42-0.70 | <0.0001 |
| 4 | 265 | 0.69 | 0.53-0.90 | 0.007 |

Models (predictors):
1; Plasma ACE2 levels.
2; Plasma ACE2 levels, age, body mass index (BMI).
3; Plasma ACE2 levels, age, body mass index (BMI), pre-existing hypertension, pre-existing heart disease, pre-existing lung disease, pre-existing kidney disease, pre-existing diabetes, pre-existing immunosuppressive condition.
4; Plasma ACE2 levels, C-reactive protein (CRP), absolute neutrophil count, and D-dimer.
n=number of patients included in statistical analysis. OR=Odds ratio for a lower (worse) WHOMax category per unit increase in plasma ACE2 level, 95%-CI=95% confidence interval, p=p-value.

**Fig. 2.**
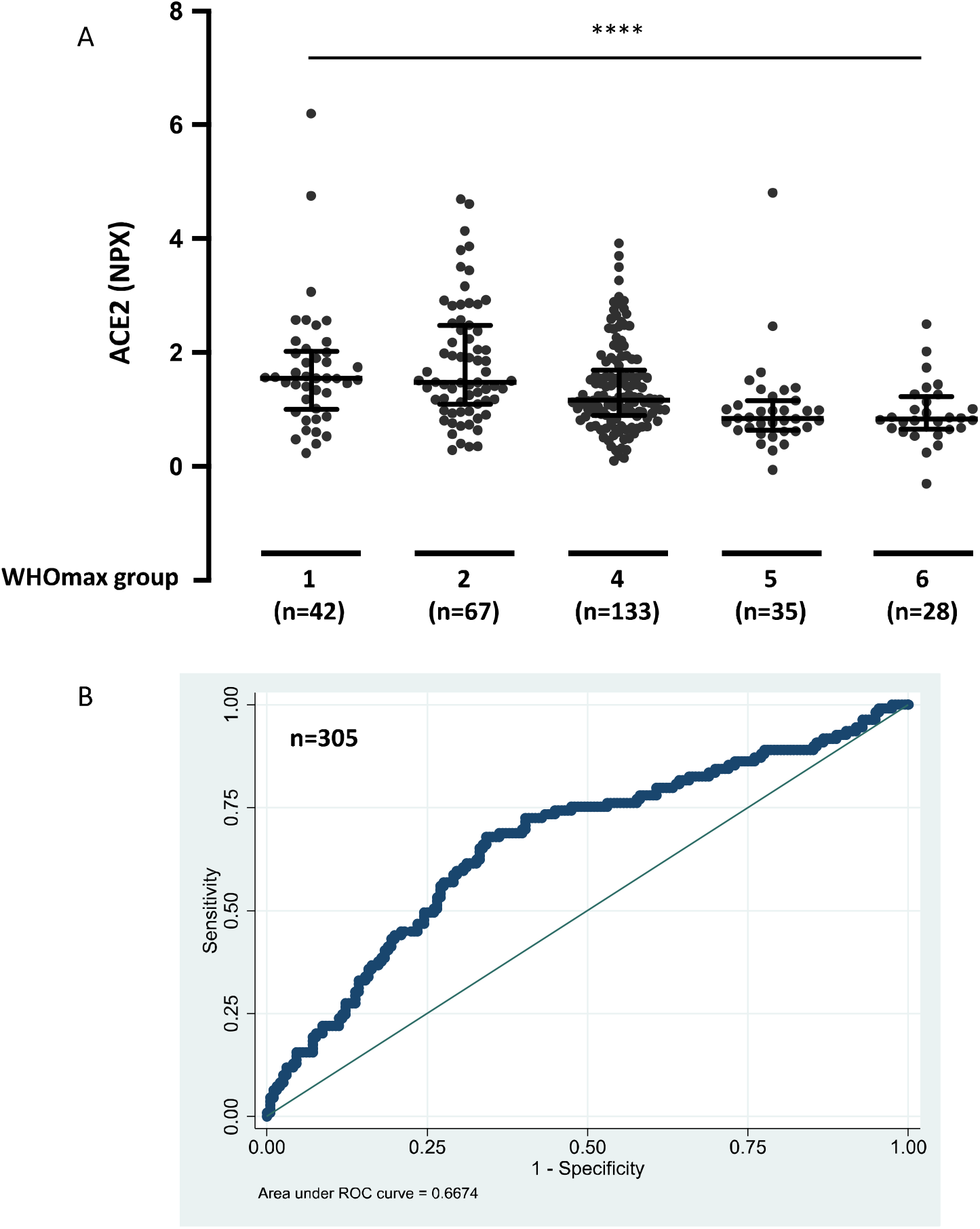
Day 0 plasma concentrations of ACE2 in COVID-19 positive patients by WHOmax groups and ROC curve. **A**. WHOmax category at 28 days is the maximum WHO score within first 28 days with death being the maximum possible. 1 = Death within 28 days. 2 = Intubated, ventilated, survived to 28 days. 4 = Hospitalized, supplementary O_2_ required. 5 = Hospitalized, no supplementary O2 required. 6 = Not hospitalized. Data were analyzed using the Kruskal-Wallis test. Bars indicate median and interquartile range. **** P < 0.0001. **B**. Receiver operating characteristic (ROC) curve comparing WHOmax groups 1-2 (died or intubated during the 28-day period) and WHOmax groups 4-6 (never intubated).

### Association between plasma ACE2 at day 0, day 3, and day 7 and WHO groups 2 (intubated and survived) and 4-5 (not intubated) in hospitalized COVID-19 patients

Next, we tested whether ACE2 plasma concentrations at day 0, day 3, and day 7 were associated with clinical status at the time of blood sampling. The patients were grouped according to WHO categories 2 (intubated and survived) or 4-5 (not intubated). High levels of ACE2 in plasma from COVID-19 patients were significantly associated with worse WHO category at the time of blood sampling at both day 0, day 3, and day 7 (*P* = 0.0004, *P* < 0.0001, and *P* < 0.0001, respectively) (Fig. 3).

**Fig. 3.**
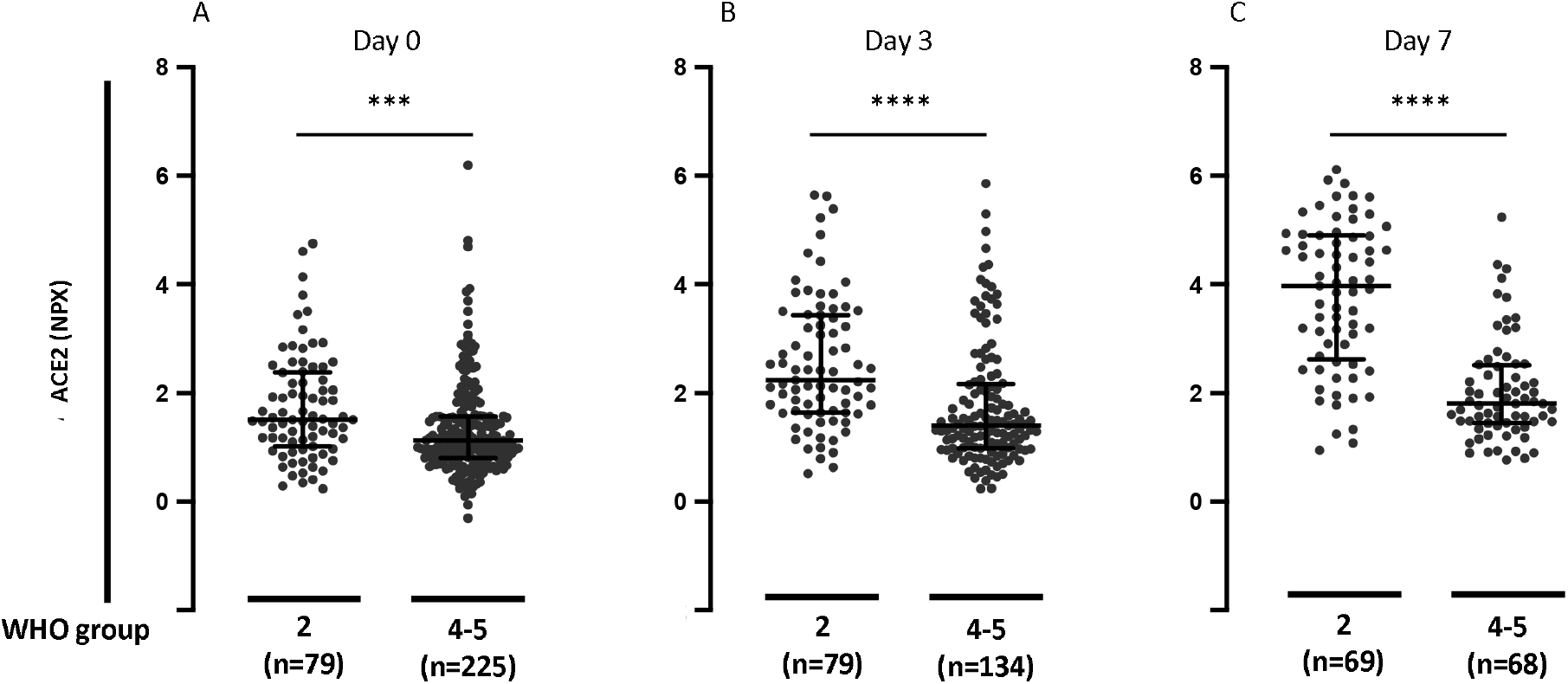
Plasma concentrations of ACE2 in hospitalized COVID-19 positive patients divided in WHO groups 2 (intubated) or 4-5 (not intubated). **A**. Day 0 plasma concentrations of ACE2 in hospitalized COVID-19 positive patients divided in WHO groups 2 (intubated) or 4-5 (not intubated) for day 0 study window (enrollment plus 24 hours). **B**. Day 3 plasma concentrations of ACE2 in hospitalized COVID-19 positive patients divided in WHO groups 2 (intubated) or 4-5 (not intubated) for day 3 study window. **C**. Day 7 plasma concentrations of ACE2 in hospitalized COVID-19 positive patients divided in WHO groups 2 (intubated) or 4-5 (not intubated) for day 7 study window. WHO categories: 2 = Intubated, ventilated. 4 = Hospitalized, supplementary O2 required. 5 = Hospitalized, no supplementary O2 required. Bars indicate median and interquartile range. Data were analyzed using the Mann Whitney test. *** P< 0.001. **** P < 0.0001.

### Association between plasma ACE2 and comorbidities in COVID-19 patients

We then analyzed the relation between ACE2 levels and comorbidities. The levels of ACE2 in plasma from COVID-19 positive patients with hypertension were significantly increased compared with plasma from patients without hypertension (*P* = 0.0045) (Fig. 4). The plasma ACE2 levels were also significantly increased in patients with pre-existing heart conditions and in patients with pre-existing kidney disease compared with patients without these pre-existing conditions (*P* = 0.0363 and *P* = 0.0303, respectively) (Fig. 4). There was no significant difference in plasma ACE2 levels comparing patients with or without pre-existing lung disease, diabetes, or immunosuppressive conditions (*P* = 0.953, *P* = 0.291, and *P* = 0.237, respectively) (Fig. 4).

**Fig. 4.**
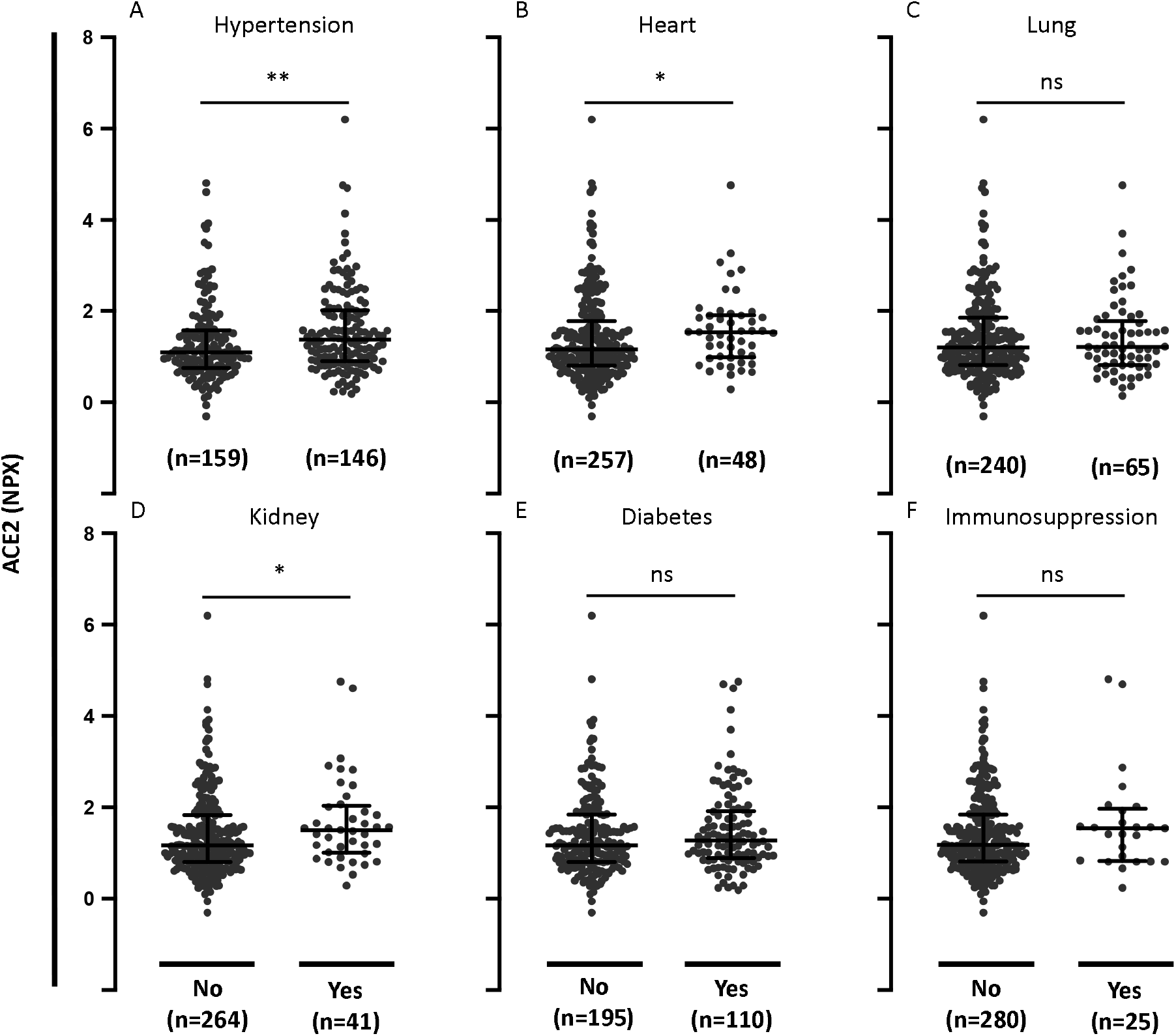
Plasma concentrations of ACE2 in COVID-19 positive patients with comorbidities. **A**. Plasma concentrations of ACE2 in COVID-19 positive patients with or without pre-existing hypertension. **B**. Plasma concentrations of ACE2 in COVID-19 positive patients with or without pre-existing heart disease (coronary artery disease, congestive heart failure, valvular disease). **C**. Plasma concentrations of ACE2 in COVID-19 positive patients with or without pre-existing lung disease (asthma, COPD, requiring home O_2_, any chronic lung condition). **D**. Plasma concentrations of ACE2 in COVID-19 positive patients with or without pre-existing kidney disease (chronic kidney disease, baseline creatinine >1.5, ESRD). **E**. Plasma concentrations of ACE2 in COVID-19 positive patients with or without pre-existing diabetes (pre-diabetes, insulin and non-insulin dependent diabetes). **F**. Plasma concentrations of ACE2 in COVID-19 positive patients with or without pre-existing immunocompromised condition (active cancer, chemotherapy, transplant, immunosuppressant agents, asplenic). Data were analyzed using the Mann Whitney test. Bars indicate median and interquartile range. * P< 0.05. ** P< 0.01. ns = not significant.

### Association between plasma ACE2 and age and BMI in COVID-19 patients

We further tested associations between ACE2 levels and age and body mass index (BMI). The levels of ACE2 in plasma from COVID-19 positive patients significantly increased with increasing age (*P* = 0.0001) (Fig. 5). There was no significant association between plasma ACE2 levels and BMI (*P* = 0.497) (Fig. 5).

**Fig. 5.**
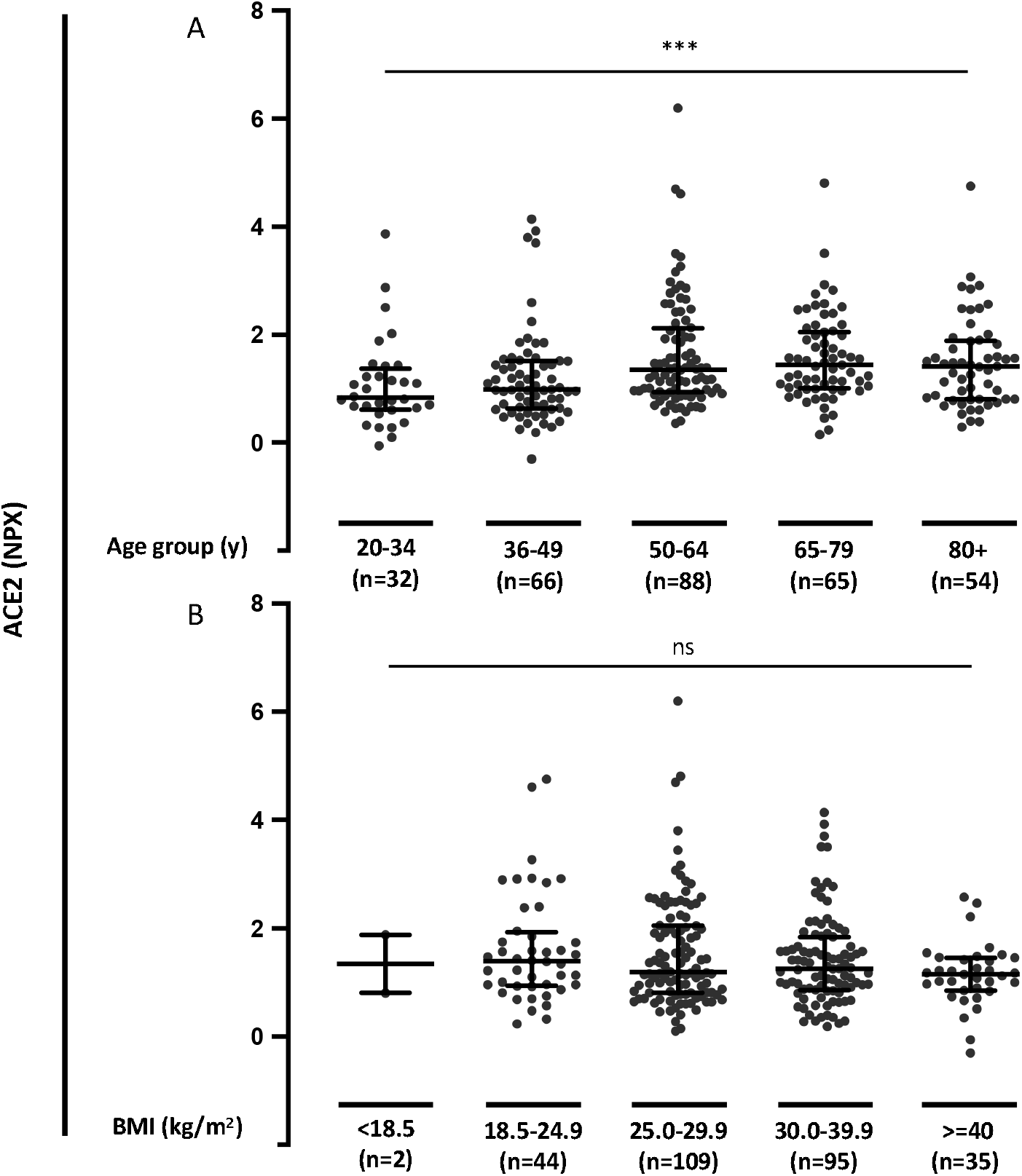
Plasma concentrations of ACE2 in COVID-19 positive patients in relation to age and BMI. **A**. Plasma concentrations of ACE2 in COVID-19 positive patients divided into age groups. **B**. Plasma concentrations of ACE2 in COVID-19 positive patients divided into body mass index (BMI) groups. Data were analyzed using the Kruskal-Wallis test. *** P< 0.001. ns = not significant.

### Association between day 0 plasma ACE2 and WHO groups 1-2 and 4-6 in COVID-19 positive and negative patients

Next, we analyzed whether ACE2 levels were different in COVID-19 patients compared with patients with respiratory symptoms without COVID-19. The levels of ACE2 showed a clear overlap between the two groups. There was no significant difference between the plasma ACE2 levels in COVID-19 positive and negative patients (*P* = 0.13). Finally, we tested whether the association between ACE2 levels and the maximum WHO score within the first 28 days were found in both COVID-19 positive and negative patients. The patients were grouped according to WHO categories 1-2 (died or intubated during the 28-day period) or 4-6 (hospitalized without intubation or not hospitalized). High levels of ACE2 in plasma from COVID-19 positive patients were significantly associated with worse WHOmax category at 28 days (*P* < 0.0001) (Fig. 6). There was no significant association between ACE2 levels and WHOmax category in COVID-19 negative patients (*P* = 0.085) (Fig. 6). To examine whether ACE2 levels were differentially distributed in WHOmax groups in COVID-19 positive vs negative patients, we also conducted a two-way analysis of variance (ANOVA). High levels of ACE2 were significantly associated with worse WHOmax category during 28 days in plasma from COVID-19 positive patients and not in COVID-19 negative patients. However, differences were not statistically different (Table 2).

**Table 2.** Associations between day 0 plasma concentrations of ACE2 and WHOmax group in COVID-19 positive and negative patients in two-way analysis of variance (ANOVA).

|  | WHO group 1-2 - 4-6 | p | 95%-CI |
| --- | --- | --- | --- |
| COVID-19 negative | 0.33 | 0.15 | -0.12 - 0.77 |
| COVID-19 positive | 0.49 | <0.0001 | 0.27 - 0.71 |
| Difference | 0.16 | 0.52 | -0.34 - 0.66 |

WHOMax category at 28 days is the maximum WHO score within first 28 days with death being the maximum possible.
p=p-value, 95%-CI=95% confidence interval.

**Fig. 6.**
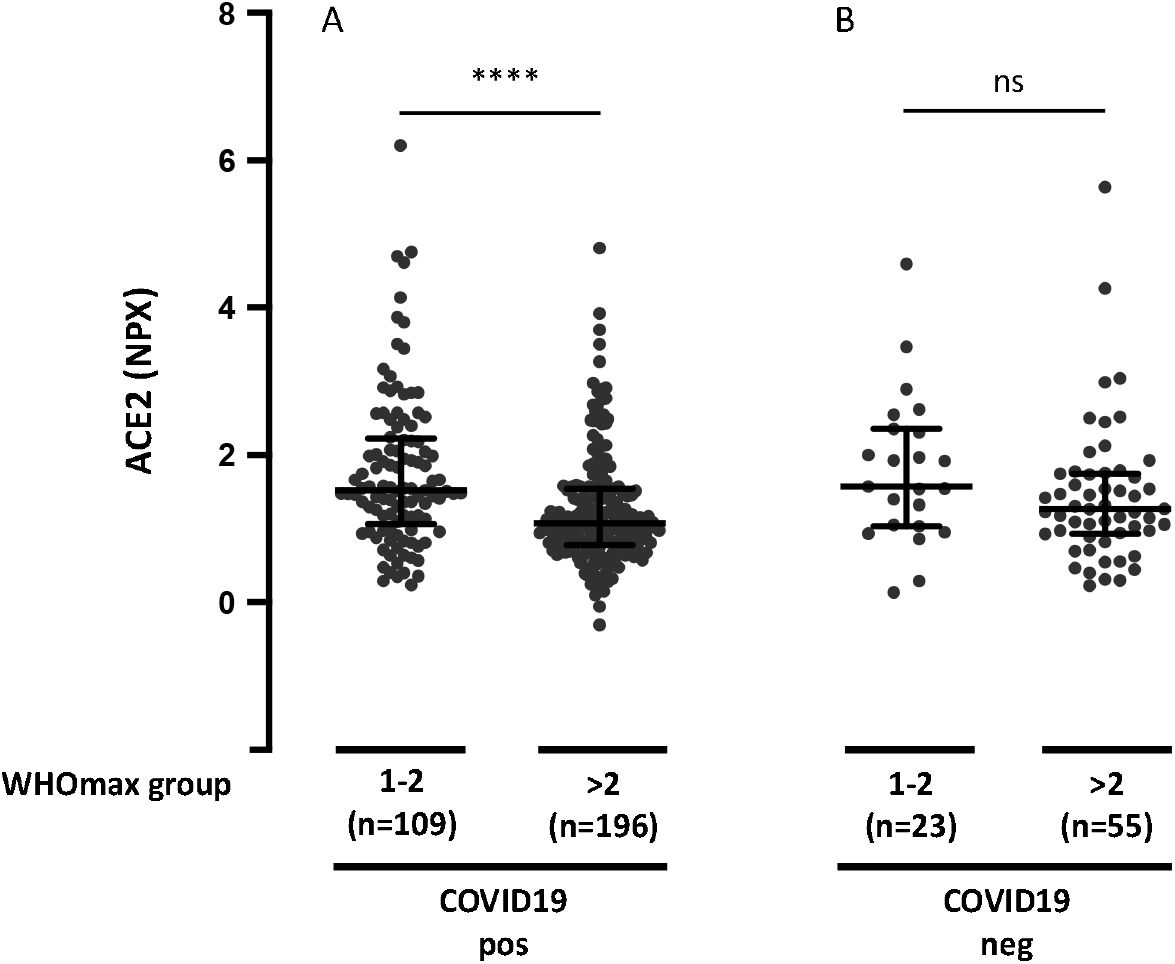
The ACE2 plasma concentration and WHOmax group in COVID-19 positive and negative patients. **A**. The ACE2 plasma concentration and WHOmax group in COVID-19 positive patients. **B**. The ACE2 plasma concentration and WHOmax group in COVID-19 negative patients. WHOmax group is the maximum WHO score within first 28 days with death being the maximum possible. 1 = Death within 28 days. 2 = Intubated, ventilated, survived to 28 days. 3 = Non-invasive ventilation or high-flow nasal cannula (not used in the treatment of COVID-19 or suspected COVID-19). 4 = Hospitalized, supplementary O2 required. 5 = Hospitalized, no supplementary O2 required. 6 = Not hospitalized. Data were analyzed using the Mann Whitney test. Bars indicate median and interquartile range. **** *P* < 0.0001. ns = not significant.

## Discussion

SARS-CoV-2 uses ACE2 as a functional receptor for entry into cells.^22^ ACE2 levels are elevated in serum from men and persons with comorbidities predicting worse COVID-19 outcome.^15,17^ This is the first description of ACE2 plasma levels in association with disease outcomes in patients with COVID-19 disease. Data were retrieved from a publicly available dataset from a large longitudinal COVID-19 study with clinical data including WHO category-based outcome.^19^

High baseline levels of ACE2 in plasma from COVID-19 patients were significantly associated with worse WHOmax category during 28 days. This indicates that abundant ACE2 expression could be involved in increased viral spread and disease burden, as previously shown in experimental models of SARS-CoV infection,^22,23^ with similar mechanisms postulated to be relevant for SARS-CoV-2 infection.^24^ The association of high ACE2 serum levels with worse clinical outcomes would in theory support blockade of ACE2 as a therapeutic strategy. In fact, ACE2 is highly expressed in the type II alveolar pneumocytes,^9^ which produce surfactant and act as progenitors for the type I alveolar pneumocytes^25^. The binding of the virus to the ACE2 on the type II alveolar pneumocytes leads to depletion of these cells, with deficit in the production and secretion of surfactant, as wells as a lack of ability to regenerate and repair injured lung tissue, leading to the exacerbation of lung injury in severe COVID-19.^25,26^ This interpretation is however corroborated by findings in animal models, showing how ACE2 protects from severe acute lung injury.^27^ Induction of severe acute lung injury through acid aspiration, ACE2-knockout mice had worsened oxygenation, massive pulmonary edema, and increased inflammatory cell infiltration compared to the wild type mice. In wild type mice, acid aspiration provoked a marked downregulation of ACE2 and an increase in Ang II levels without affecting ACE levels. In knockout mice, Ang II levels increased to a greater extent, promoting further lung damage. Moreover, Ang II has been found to activate several cells within the immune system, such as macrophages, leading to a higher production of proinflammatory cytokines like IL-6 and TNFα.^25,26,28^ Therefore, the direct blockade of ACE2, by removing the physiological brakes on the angiotensin system, could have detrimental effects on COVID-19 disease. For these reasons, an alternative strategy to inhibit the interaction of SARS-CoV-2 with ACE2 could be the administration of recombinant soluble ACE2 as a decoy receptor, which has been previously tested in small clinical trials in acute respiratory distress syndrome^29^ and is now being explored in COVID-19 disease.^30^ However, the level of ACE2 in the plasma does not necesarily reflect the expression of ACE2 in the plasma membrane of host cells. Thus, high levels of ACE2 in the plasma might result from increased lysis of ACE2-expressing cells as a consequence of more severe infection.

It is desirable to predict disease outcomes in order to specialize treatment, since the drugs used in the treatment of COVID-19 can have deleterious side effects.^31^ The ROC curve showed moderate value of measuring baseline ACE2 for WHOmax groups 1-2 (died or intubated during the 28-day period) vs WHOmax groups 4-6 (hospitalized without intubation or not hospitalized). Further, differences in ACE2 plasma levels between patients in WHO categories 2 (intubated and survived) vs 4-5 (not intubated) were more evident on day 3 and day 7 of admission. Therefore, repeated analysis of ACE 2 levels could increase the predictive value. This is in line with a recent study showing increasing SARS-CoV-2 during hospitalization and prolonged viral shedding in more severe COVI-19 disease.^32^ However, it is important to recognise that many patients did not have samples drawn on day 3 and/or day 7. This was primarily because patients had been discharged from hospital. The population sampled on day 3 and day 7 therefore consists of patients with more severe disease compared with day 0. Anyway, it is interesting that the association between levels of ACE2 and WHOmax category during 28 days in plasma from COVID-19 patients was statistically significant also after correction for age, BMI, pre-existing medical conditions, and the laboratory test CRP, absolute neutrophile count and D-dimer. This means that plasma ACE2 levels can add to the value of already existing data to help predict disease outcome in COVID-19. ACE2 is a membrane-bound enzyme, and therefore measuring the circulating and urine levels of ACE2 is complex.^28^ Soluble ACE2 is the result of the cleavage of the membrane-bound ACE2 by disintegrin and metalloprotease 17. Interestingly, there is a correlation between plasma ACE activity and ACE2 activity in healthy individuals.^33^ If this same correlation is seen in patients with COVID-19, plasma ACE levels could be used to approximate the activity of ACE2 in COVID-19 positive patients. This is noteworthy because serum ACE analysis is a standardized test in most international hospitals.

The levels of ACE2 in plasma from COVID-19 positive patients with hypertension were significantly increased compared with plasma from patients without hypertension. Patients with hypertension are often treated with ACE-inhibitors and AT1R-blockers. During this pandemic, it has been highly debated whether or not the use of these antihypertensive medications should be discontinued in patients with COVID-19. A study performed on rats showed that the use of ACE inhibitors and/or use of AT1R-blocker led to an increased expression of ACE2.^34^ This has raised the concern that patients with hypertension, who are treated with ACE2-modulating drugs like ACE-inhibitors or AT1R-blockers might be at a higher risk for severe COVID-19 infection, since it could increase the entry-way for the virus.^35^ A recent study showed that plasma ACE2 activity is increased in COVID-19 patients treated with ACE inhibitors.^36^ However, more severe COVID-19 disease in patients treated with ACE-inhibitors or AT1R-blockers has not been supported by recent population based data.^37^ The plasma ACE2 levels were also significantly increased in patients with pre-existing heart conditions and in patients with pre-existing kidney disease. This is in line with the central role of RAAS in these conditions. In contrast, ACE2 levels does not seem to explain the risk of severe COVID-19 disease associated with pre-existing lung disease, diabetes, or immunosuppression. Plasma ACE2 was associated with age, which is in line with recent observations showing higher serum levels of ACE2 in adults compared to a pediatric cohort.^38^ There were no associations between plasma ACE2 levels and BMI.

Baseline ACE2 levels did not show statistically significant differences between COVID-19 positive and negative patients, which is in line with the results of a recent study with a much smaller sample size.^39^ The associations between high plasma levels of ACE2 and worse WHOmax category during 28 days were more pronounced in COVID-19 positive patients compared with COVID-19 negative patients but the difference was not significant in the two-way ANOVA analysis. It is therefore not possible with this dataset to conclude whether differences in ACE2 levels are specifically associated with disease outcome in patients with COVID-19 positive disease only.

There are some overall limitations with this study. First, the plasma ACE2 levels were measured as relative protein concentrations using NPX (Normalized Protein eXpression) values. Therefore, it is not possible to determine a plasma ACE2 cut-off value to predict severe outcome. Second, several continuous variables were categorized in the publicly available data set. This decreases the statistical power of some of the analysis. Third, some potentially important variables such as gender and treatment were not available. Finally, obviously causality of associations between plasma ACE2 levels and severity of COVID-19 disease cannot be concluded from this study.

Overall, this study suggests a potential role of measuring ACE2 in COVID-19 to predict disease outcome. Further, ACE2 levels could be a link between severe COVID-19 disease and its risk factors, namely hypertension, pre-existing heart disease and pre-existing kidney disease. The design of the data analysis using the Olink platform does not allow assessment of quantitative differences. However, previous studies have described a positive correlation between plasma ACE2 and ACE1 activity. This is interesting because ACE1 (serum ACE) analysis is a standardized test in most hospital laboratories. Therefore, our study encourages quantitative investigations of both plasma ACE 1 and 2 in COVID-19.

## Data Availability

Data is publicly available.

https://www.olink.com/mgh-covid-study/

## Abbreviations

ACE: Angiotensin converting enzyme
Ang I: Angiotensin I
Ang II: Angiotensin II
ANOVA: Analysis of variance
AT1R: Type 1 angiotensin II receptor
AUC: Area under the curve
BMI: Body mass index
CoV: Coronavirus
COVID-19: Corona virus
disease 2019 CRP: C-reactive protein
IL: Interleukin
NPX: Normalized Protein eXpression
PEA: Proximity extension assay
RAAS: Renin-angiotensin-aldosterone-system
ROC: receiver operating characteristic
SARS-CoV-2: Severe acute respiratory syndrome coronavirus 2
TNF: Tumor necrosis factor

## Authors’ contributions

MF, AM, NH, and MG organized enrollment and processing of the prospective study. IG was involved in the Olink analyses. TWK initiated the project concerning ACE2. PQ and ALLN extracted the ACE2 data from the publicly available data set. TWK and BMB made the statistical analyses. TWK, HSS and FR drafted the first version of the manuscript. All authors were involved in interpreting the data, revising the manuscript and approved the final submission.

## Acknowledgements

Data provided by the MGH Emergency Department COVID-19 Cohort (Filbin, Goldberg, Hacohen) with Olink Proteomics. We want to thank Aparna Udupi, Department of Biostatistics, Aarhus University for technical assistance running the statistical analyses.

## Ethics

Sample collection and analysis was approved by Partners Human Research Committee (PHRC).

